# Long-term positive effects of olfactory training on quality of life and subjective measures of olfactory function

**DOI:** 10.64898/2025.12.09.25341911

**Authors:** Anja L. Winter, Pernilla Sahlstrand Johnson, Johan N. Lundström

**Author notes:** CORRESPONDING AUTHORS Anja L. Winter, Department of Clinical Neuroscience, Nobels väg 9, 171 77 Stockholm, Sweden; Johan N. Lundström, Department of Clinical Neuroscience Karolinska Institutet, Karolinska Institutet, Nobels väg 9, 171 77 Stockholm, Sweden.

## Abstract

Olfactory dysfunction following viral infection is a debilitating condition. Currently, the recommended treatment for olfactory dysfunction is olfactory training, a systematic and repeated exposure to odors over a prolonged period. A new tool has been proposed for olfactory training that present odors using nasal inserts rather than hand-held devices, to increase ease of training and thereby improving compliance and adherence to the training regimen. Here, we aimed to determine how olfactory training affects subjective experience of quality of life and olfactory function and potential differences training using regular versus the nasal insert setup. Using a randomized controlled design (*N* = 111), we investigated the effects of an 8-week olfactory training regimen using nasal inserts compared to standard care olfactory training in individuals with post-viral functional hyposmia. Subjective measures of quality of life as well as qualitative and quantitative olfactory function were assessed before and after the training regimen, and during a 1-year follow-up. Overall, participants showed significant and sustained improvements in both subjective olfactory function and quality of life following training. Critically, the nasal insert group demonstrated greater gains in social functioning and quantitative olfactory scores shortly after training, with enhanced olfactory benefits persisting at the 1-year follow-up. The magnitude of changes in quality of life was correlated with subjective olfactory improvement overall, with a stronger association for the nasal insert group. Thus, nasal insert training appears to enhance perceived olfactory function, which coincides with improved quality of life. Findings from this trial provide insight into the benefits of olfactory training on subjective functioning and quality of life, as well as the efficacy of nasal insert training in post-viral hyposmia.

---

The sense of smell plays a fundamental role in human health and wellbeing given that it guides and affects nutrition, safety, interpersonal interactions, as well as various types of emotional experiences (Boesveldt & Parma, 2021). Losing olfactory functions therefore often result in significant distress with considerable implications for daily life and psychological wellbeing. Olfactory dysfunction following viral infections has after the COVID-19 pandemic gained renewed clinical and public attention due to its prevalence and substantial impact on quality of life (Jaime-Lara et al., 2020). In fact, common viral infections of the upper respiratory tract are one of the most frequent causes of olfactory dysfunction (Seiden, 2004) and individuals with smell loss frequently report decreased appetite, impaired detection of environmental hazards (such as gas leaks or spoiled food), reduced enjoyment of eating and social activities, and a worry related to coping with long-term dysfunctions; all contributing to a lower quality of life (Croy et al., 2014).

Among the available interventions for olfactory dysfunction, olfactory training (often referred to as ‘smell training’) has become the most widely recommended approach for promoting olfactory recovery. This therapeutic method is aimed to enhance regeneration and reorganization within the olfactory system by a structured and repeated exposure to specific odorants over an extended period of time (Thomas Hummel et al., 2009). Clinical studies have shown that olfactory training leads to measurable improvements in olfactory threshold, discrimination, and identification ability in post-infectious olfactory loss (Damm et al., 2014). Despite the treatment being relatively efficient, individuals who are prescribed it rarely complete the full regimen, resulting in a smaller number than optimal benefiting from the training. Although precise data on drop-out rates and protocol deviation relating to olfactory training are lacking, non-adherence rates for comparable interventions reach as high as 50% for chronic medication and 70% for physiotherapy (DiMatteo et al., 2002; Sluijs et al., 1993). To address these limitations and the known challenges associated with adherence to the olfactory training regimen, a new tool for continuous and effortless odor delivery through the use of nasal inserts containing odorants was recently validated (A L Winter et al., 2025). Compared to standard care olfactory training using hand-held odor delivery devices, often jars, this approach had similar or better results in objective measures of olfactory function and significantly improved the adherence of patients engaging in smell training, increasing it to a 93% compliance rate.

In the present study, we aimed to investigate not only the 1-year long-term effects of olfactory training on participants’ subjective perception of their qualitative and quantitative smell function but also its broader impact on quality of life. Beyond assessing these outcomes, we sought to compare the effectiveness of conventional standard care olfactory training, as presently recommended by the Swedish health care system, with that of a nasal insert-based approach, designed to simplify and enhance adherence to treatment. By exploring whether this alternative delivery method can offer comparable or improved therapeutic benefits, we aimed to shed new light on how olfactory rehabilitation can be made more accessible and effective for individuals struggling with olfactory dysfunction and related declines in quality of life.

## METHOD

### Participants

Participants (*N* = 173) were recruited from two outpatient clinics and via social media advertisement. Inclusion criteria were post-viral functional hyposmia at baseline. All participants were assessed using the clinically validated Sniffin Sticks test, assessing odor detection threshold (T), odor quality discrimination (D), and cued odor quality identification (I); subsequently summed into a TDI score (T Hummel et al., 1997). To qualify for the study, participants had to be between 18 and 65 years old and have a TDI score between 15.25 and 31.25 at baseline. Exclusion criteria were psychiatric diagnoses, non-viral causes of olfactory dysfunction (such as head trauma, sinonasal disease, surgery, etc.), and current enrollment in other olfactory training studies. After initial screening and excluding those with a TDI score outside our preregistered inclusion criteria (*n* = 50), a total of 123 participants were enrolled in the study. Twelve participants were subsequently excluded before analysis: 4 due to problems with testing conditions, 2 due to nasal congestion, 1 due to not being able to attend the second visit, and 5 due to missing baseline questionnaire data. The final sample reported in this manuscript consists of 111 individuals. Study procedures were in accordance with the Declaration of Helsinki, approved by the Swedish Ethical Review Authority (Dnr: 2023-03779-01), and all participants provided written informed consent prior to participation.

### Procedure

Participation entailed two visits, one at baseline and one following an 8-week olfactory training regimen, as well as a 1-year follow-up consisting of online questionnaires. At the baseline visit, quality of life was assessed, as well as subjective and objective olfactory function (A L Winter et al., 2025). Information on participant demographics was also collected, see Table 1. Participants were randomly assigned to an olfactory training regimen using either nasal inserts (n = 59) or standard care (n = 52). The post-training visit mirrored the baseline visit with the exception that questions about demographics were replaced with questions regarding adherence to the training protocol (A L Winter et al., 2025).

**Table 1.** Descriptive statistics of research participants collected during baseline visit.

|  | Nasal Insert<br>(N = 59) | Standard Care<br>(N = 52) | Total<br>(N = 111) |
| --- | --- | --- | --- |
| **Sex** |  |  |  |
| Male | 10 | 10 | 20 |
| Female | 49 | 42 | 91 |
| **Age** |  |  |  |
| Mean (SD) | 49.4 (12.1) | 47.6 (8.65) | 48.5 (10.6) |
| **TDI** |  |  |  |
| Mean (SD) | 24.9 (4.61) | 24.9 (4.69) | 24.9 (4.63) |
| **Dysfunction duration (months)** |  |  |  |
| Mean (SD) | 42.9 (41.6) | 43.7 (48.7) | 43.3 (44.9) |

#### Nasal Insert (NI)

Participants received single use scented nasal inserts (Nosaplugs, NosaMed AB, Stockholm; Figure 1A) to wear for 20 minutes in the morning and 20 minutes in the evening during every weekday for 8 straight weeks. Weekends were designated rest days without training to allow the olfactory system time of reduced stimulation. The nasal inserts are placed into the nostrils (Figure 1B) and administer one specific scent amongst 10 (vanilla, lemon, melon, rosemary, menthol, orange, peach, strawberry, cherry, or cola) while allowing near normal nasal patency. Each session used two odors (10 minutes each), totaling four different scents per day. To assist in dispersing the different scents throughout the training regimen, a schedule of which odor to use was provided. Participants were instructed to focus on the smell and to discard the plugs after use.

**Figure 1.**
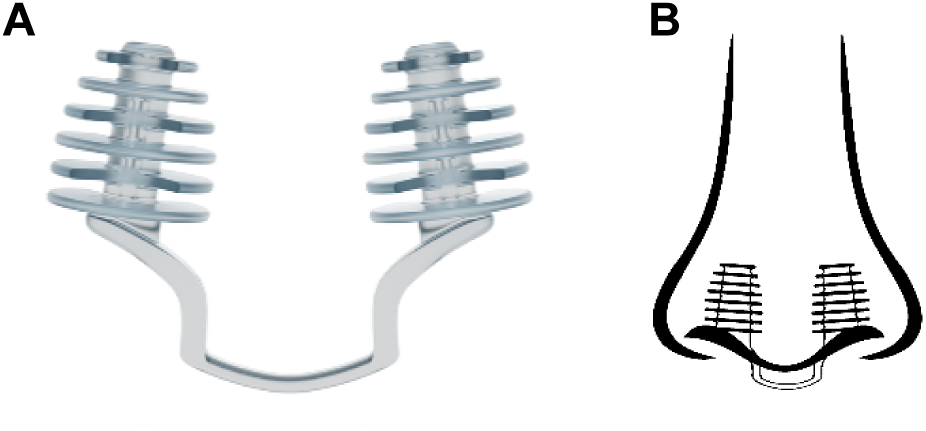
A. Image of the nasal insert with its scented lamellae that allow near normal nasal patency. B. Schematic drawing of a nasal insert positioned within the nose, viewed from the front.

#### Standard Care (SC)

Participants selected 4–6 household odors and were instructed to repeatedly smell each odor for 10–20 seconds, rotating among the different odors for a total of 20 minutes in the morning and 20 minutes in the evening on weekdays for 8 weeks. Participants were also instructed to focus on the smell during the sessions. In Sweden, the recommended standard olfactory training is approximately 12 weeks performed daily, which is longer than what is currently reported in the literature. Our regimen was shortened to 8 weeks and limited to weekdays to reduce the high non-compliance observed in the clinical setting. Besides these changes listed, the training regimen closely followed present national recommendations.

### Measurement

#### Subjective olfactory dysfunction

##### Quantitative

Subjective quantitative olfactory function over the past three days was self-assessed using a 10-point visual analogue scale (VAS), ranging from 0 (no sense of smell) to 10 (excellent sense of smell).

##### Qualitative

Subjective qualitative olfactory function was also reported by participants answering whether they experienced qualitative olfactory dysfunction, in terms of phantosmia (smell hallucinations) or parosmia (distorted smells) via yes or no questions. Participants were also tasked to rate the severity of such using a 10-point visual analogue scale (VAS), ranging from 0 (very minor issues) to 10 (very significant issues).

#### Quality of life (QoL)

##### Olfactory dysfunction-related QoL

The 28-item Olfactory Dysfunction Outcomes Rating (ODOR-28) was used as a measure of olfactory dysfunction-related QoL. ODOR-28 is a validated (Lee et al., 2022) self-report questionnaire measuring olfactory dysfunction-related effects on quality of life. The assessment contains 28 questions that participants scored from 1 (very rarely bothered) to 5 (very frequently bothered), yielding a total score that ranged from 28 to 140, with higher scores indicating lower quality of life.

##### Mental and physical health-related QoL

The Short Form 36 Health Survey (SF-36) was used to measure mental and physical health-related quality of life. SF-36 is a validated (Brazier et al., 1992) self-report instrument with broad applicability that assesses mental and physical health, with a focus on physical health. The different items contain various questions and alternatives. Scores were recoded and summarized in accordance with the scoring rules for the RAND 36-Item Health Survey (Version 1.0) (Ware & Sherbourne, 1992), which yields a total of eight scores across the following subscales; physical functioning, role limitations due to physical health, pain, general health, vitality, mental health, social functioning, and role limitations due to emotional problems. For all subscales, a high score defines a more favorable state of health.

### Statistical analysis

Due to a number of dropouts, especially in the standard training group, we used the last observation carried forward (LOFC) method to handle the missing data points of dropouts (Alwateer et al., 2024; Digitale et al., 2025). The data of 16 participants (*n*_NI_ = 4, *n*_SC_ = 12) who dropped out were carried forward from the baseline visit to the post-training visit and 1-year follow-up. The data of 23 participants who attended the post-training visit but did not complete the 1-year follow-up (*n*_NI_ = 17 *n*_SC_ = 6) were carried forward from the post-training visit to the 1-year follow-up. In effect, the LOFC method allows comparisons between conditions with differences in compliance rates but do not assume drop-out cause, i.e. drop out due to treatment success or treatment failure. The LOFC can be considered a conservative method for protecting data integrity, given that it biases analyses towards finding no effect.

All raw data and analysis scripts are available at the Open Science Framework (OSF) https://osf.io/jtbf6/overview. The study hypothesis, inclusion/exclusion criteria and analysis plan were preregistered on clinicaltrials.gov, ID NCT06142565. Note that all pre-registered questionnaires were in the end not included in the study to avoid testing fatigue among participants. Statistical analyses included t-tests to compare group means, analyses of variance (ANOVA) for examining individual and interaction effects of variables, and analyses of covariance (ANCOVA) to compare post-test means between groups while controlling for pre-test differences. This type of analysis was chosen because its approach isolates the treatment effect more accurately and aligns with EMA and FDA recommendations for analyzing between-group treatment effects (Senn, 2006; Zimmermann et al., 2019).

All analyses were performed using the statistical software R (v4.5.1) (R Core Team, 2024) and the packages car (v3.1.2; Fox & Weisberg, 2019), dplyr (v1.1.4; Wickham et al., 2023), ggplot (v3.5.1; Wickham, 2016), table1 (v1.4.3; Rich, 2023), and tidyr (v1.3.1; Wickham et al., 2024). The significance criterion for the statistical tests was set to α =0.05.

## RESULTS

### Subjective olfactory function

#### Olfactory training yield sustained improvements in subjective quantitative function, with greater gains observed in the nasal insert group

For quantitative olfactory function, we aimed to investigate how participants perceived their subjective quantitative olfactory function during the post-training visit and the 1-year follow-up compared to their baseline assessment before olfactory training. Beginning with the whole sample, we found that olfactory training improved participants’ perceived olfactory abilities, as manifested by a significant increase in rated olfactory function between the baseline and the post training visit, t(110) = 4.9, p < .001, as well as between the baseline and the 1-year follow-up assessment, t(110) = 5.8, p < .001. Assessing differences between training methods, we found that the nasal insert group expressed significantly higher subjective quantitative olfactory scores during the post-training visit compared to the standard care group, F(1, 109) = 5.77, p = .02 (Figure 2). This difference was also present during the 1-year follow-up, F(1, 109) = 5.33, p = .02.

**Figure 2.**
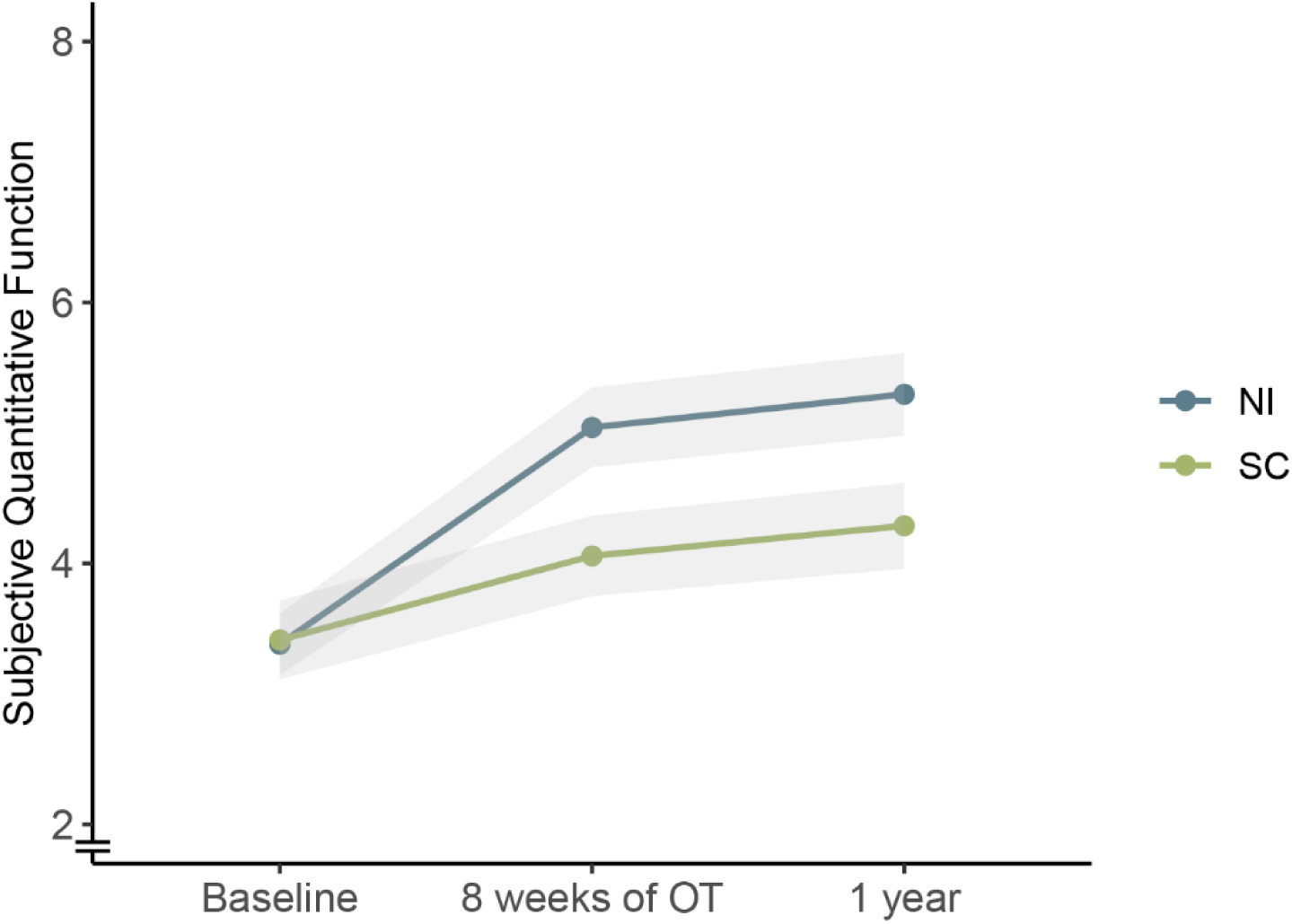
Mean subjective quantitative olfactory function scores by visit, separated by olfactory training group. Higher scores represent greater function. Shaded areas represents standard error of the mean. NI denotes training using nasal inserts and SC training according to standard care. Note break in axis.

#### Olfactory training yields sustained improvements in subjective qualitative olfactory function

For qualitative olfactory function, we first aimed to investigate the frequency of qualitative olfactory dysfunction in each group for the three different measure points, see Table 2. We then wanted to investigate how participants perceived the severity of their qualitative olfactory dysfunction during the post-training visit and the 1-year follow-up compared to their baseline assessment before olfactory training. Beginning with the whole sample, we found a significant decrease in subjective qualitative olfactory dysfunction between the baseline and the post training visit, t(110) = 4.4, p < .001, as well as between the baseline and the 1-year follow-up assessment, t(110) = 2.6, p < .001 (Figure 3). However, there was no significant difference in qualitative olfactory complaints between the nasal insert group and the standard care group.

**Figure 3.**
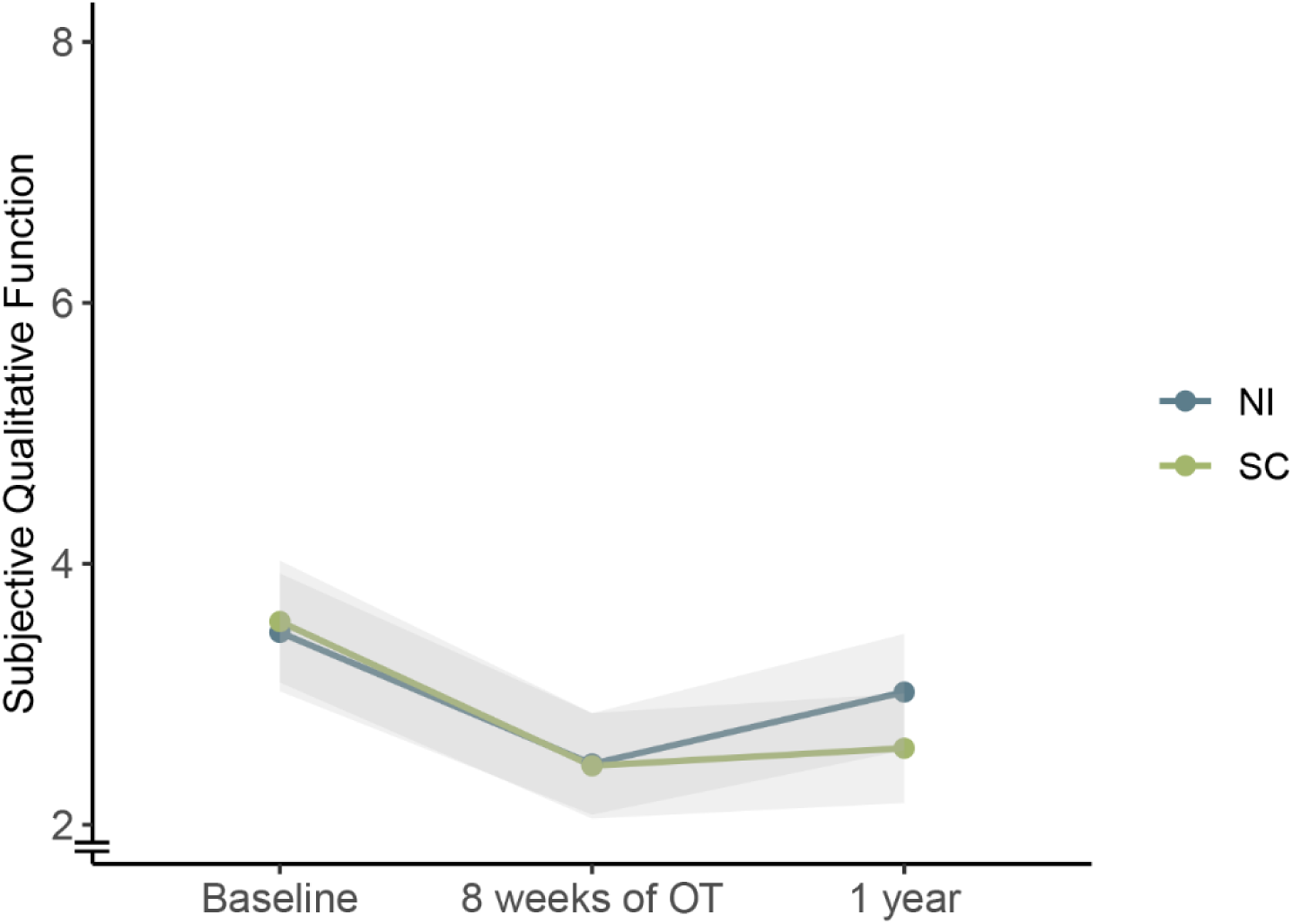
Mean subjective qualitative olfactory dysfunction scores by visit, separated by olfactory training group. Higher scores represent higher dysfunction. Shaded area represents standard error of the mean. NI denotes training using nasal inserts and SC training according to standard care. Note break in axis.

**Table 2.**
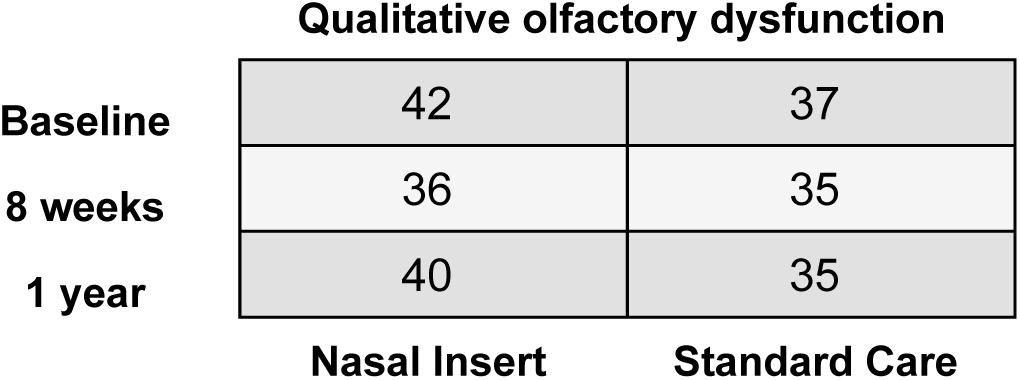
Prevalence of qualitative olfactory dysfunction in respective olfactory training group, separated by the three different measuring points.

### Quality of life (QoL)

#### Olfactory training yields sustained improvements in olfactory dysfunction-related quality of life

We then assessed the effect of olfactory training on olfactory dysfunction-related quality of life by comparing the baseline scores with the post training scores for the entire sample. We found a significant increase in olfactory dysfunction-related QoL due to olfactory training from the baseline to post training visit, t(110) = 5.04, p < .001, as well as from the baseline to the 1-year follow-up assessment, t(110) = 5.07, p < .001; Figure 4.

**Figure 4.**
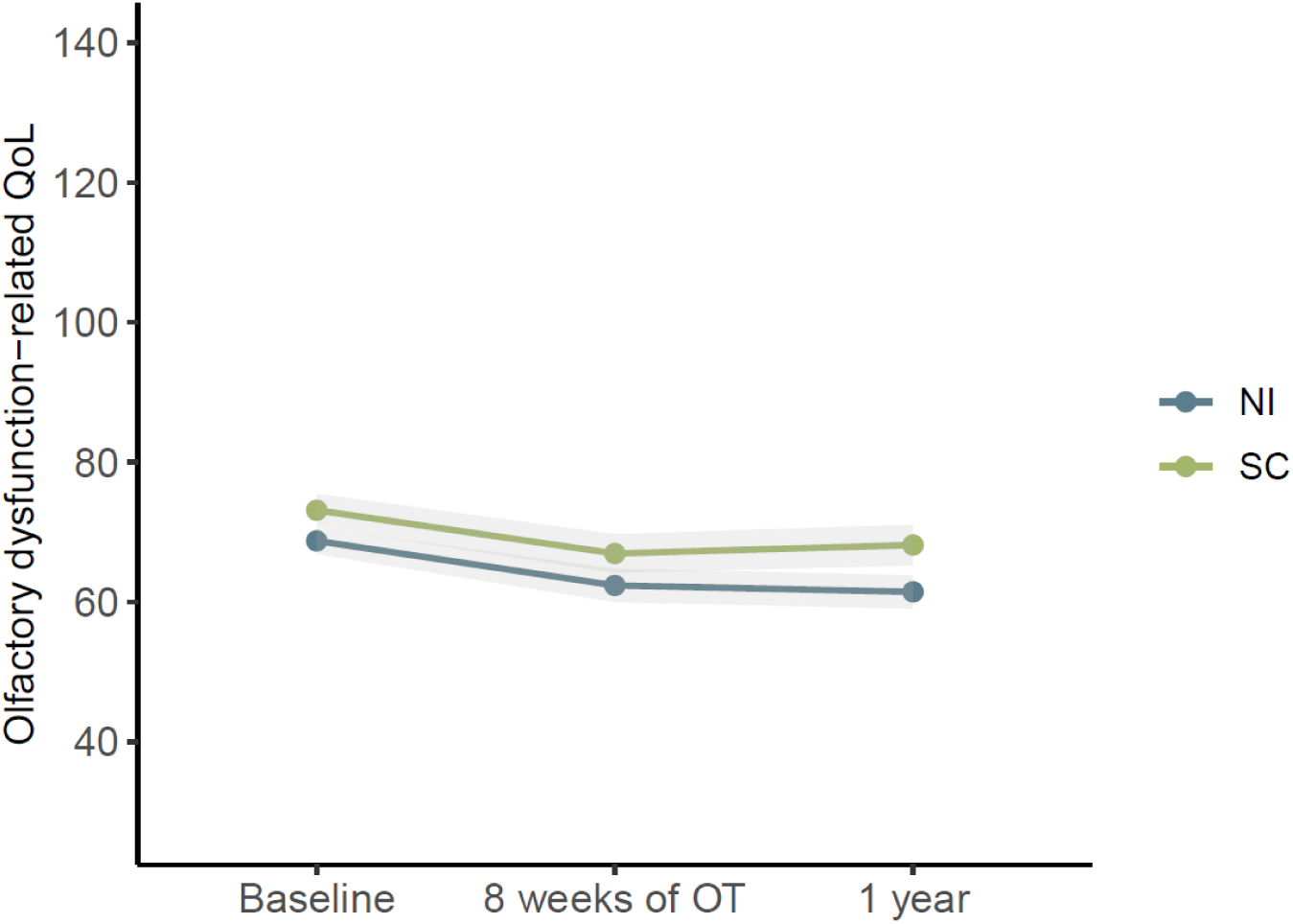
Mean olfactory dysfunction-related QoL (ODOR 28) scores by visit, separated by olfactory training group. Higher scores represent lower quality of life. Shaded area represents standard error of the mean. NI denotes training using nasal inserts and SC training according to standard care.

Next, we assessed potential differences between training groups in respect of improvement in olfactory dysfunction-related QoL. In contrast to above reported differences related to subjective experience of olfactory capabilities, where the nasal insert training group experienced better olfactory function, we found no significant difference between the groups; neither for the post training visit, nor for the 1-year follow-up. In addition, we were interested in whether the presence of qualitative olfactory dysfunction (i.e., parosmia/phantosmia) could influence the improvement in olfactory dysfunction-related QoL, either separately from or in interaction with the olfactory training group allocation (Figure 5). We found the largest nominal increase in olfactory dysfunction-related QoL for the nasal insert group without qualitative olfactory dysfunction. However, when conducting a two-way ANOVA on the effects of olfactory training group and olfactory dysfunction group on changes in ODOR-28 scores, no significant effect was found, neither in terms of main effects nor interaction effects.

**Figure 5.**
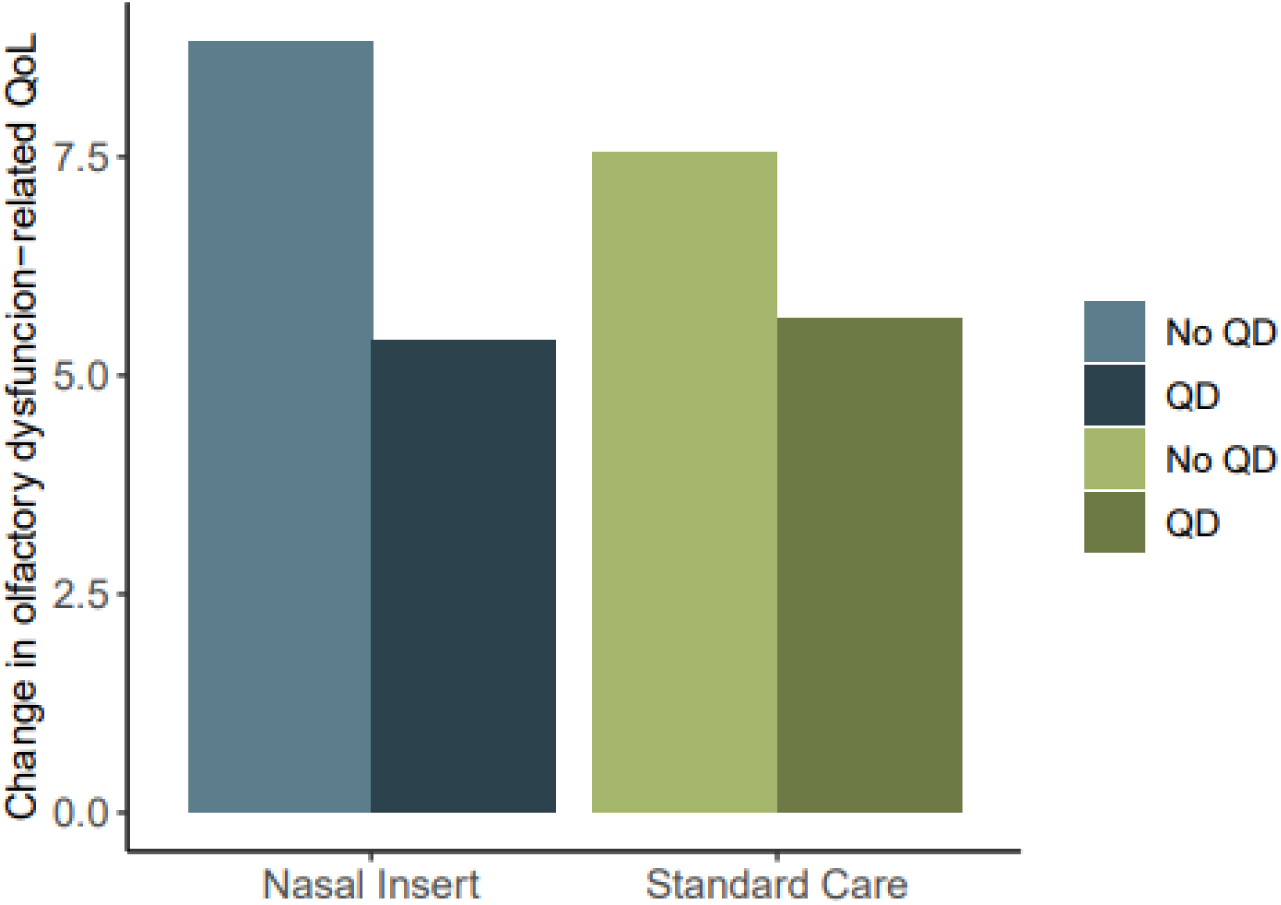
Mean changes in olfactory dysfunction-related QoL, divided by olfactory training group, separated by presence or absence of qualitative olfactory dysfunction (QD). Note that scores represent reduction in ODOR-28 scores, where greater reduction equals greater improvement in QoL.

Further, we conducted an exploratory analysis of the potential correlation between changes in subjective quantitative olfactory function and changes in ODOR-28 scores. A Spearman’s rank correlation revealed a significant negative association between the two change scores in both the nasal insert group, ρ = −.45, p < .001, and the standard care group, ρ = −.32, p < .05, indicating that increases in perceived quantitative olfactory function is correlated with decreases in olfactory dysfunction-related quality of life issues.

#### Olfactory training using nasal inserts yield greater improvement in post-training social functioning scores compared to standard care

We then investigated mental and physical health-related QoL scores during the post-training visit and the 1-year follow-up compared to their baseline assessment before olfactory training. When assessing the whole sample, we found no significant difference in any of the SF-36 subscales between the baseline and the post training visit (all t < 1.91; p > .06).

Finally, we assessed whether there was a difference between the groups on the different subtests of the SF-36, first at the post-training visit and then the 1-year follow up. The results of the analysis revealed that the nasal insert group scored significantly higher on the social functioning subscale on the post-training visit (F = 4.74, p = .03) compared to the standard care group after completed training (Figure 6). However, this difference was no longer apparent at the 1-year follow-up (F = .004, p = .09) The same type of analysis revealed no statistically significant difference between the training groups on physical functioning, role limitations due to physical health, role limitations due to emotional problems energy/fatigue, emotional well-being, or general health (all *F* < 3.31; *p* > .07).

**Figure 6.**
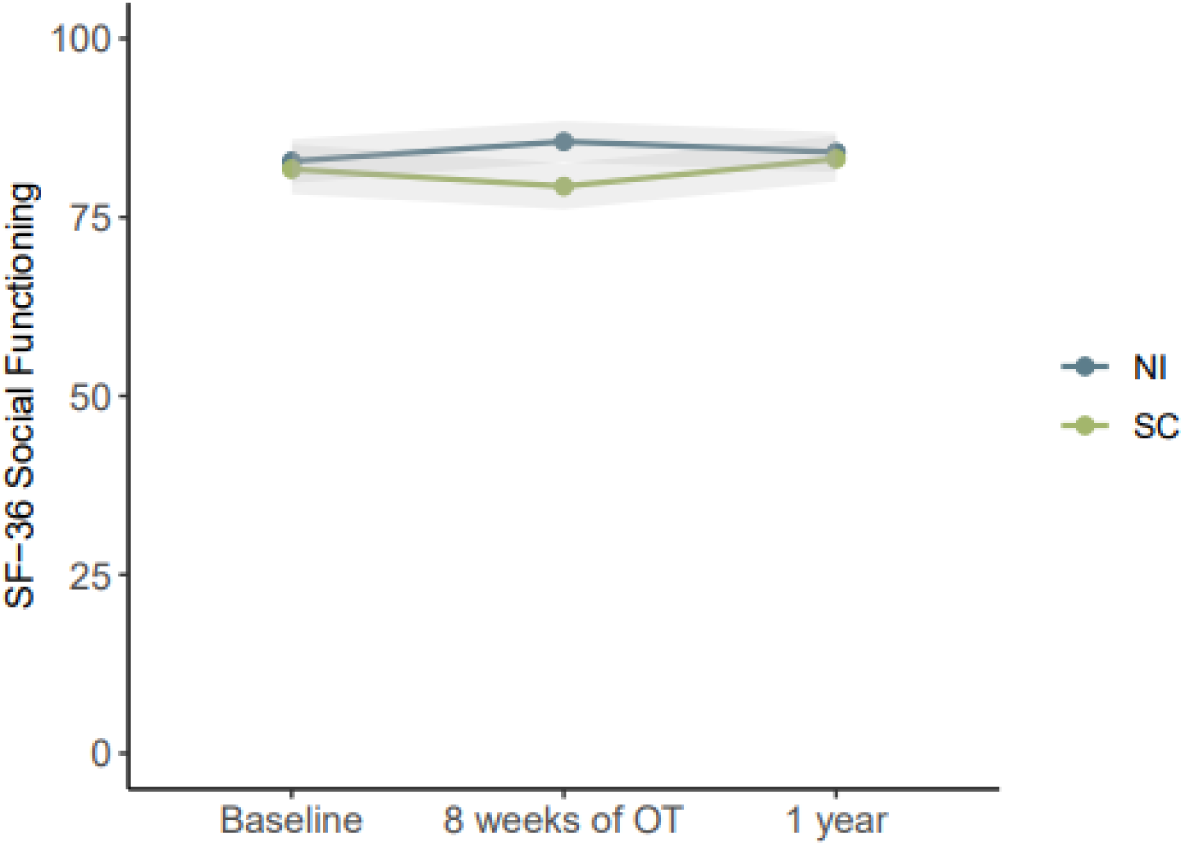
Social functioning mean scores by visit, separated by olfactory training group. Higher scores represent greater social functioning. Shaded area represents standard error of the mean. NI denotes training using nasal inserts and SC training according to standard care.

## DISCUSSION

In this study, we aimed to investigate how olfactory training could affect subjective experiences of olfactory function and quality of life on post-viral olfactory dysfunction, as well as the potential differences in these effects depending on the type of olfactory training; standard care or using the recently developed smell training technique with scented nasal inserts. Across the entire sample, participants reported a significant improvement in subjective olfactory function following training, which also was maintained at the 1-year follow-up. The nasal insert group subjectively experienced better olfactory function than the standard care group both immediately after training and at the 1-year follow-up. Participants also experienced a notable reduction in subjective qualitative olfactory dysfunction from baseline to both post-training and 1-year follow-up assessments. Olfactory dysfunction-related quality of life improved significantly after training and remained higher at the 1-year follow-up. The magnitude of subjective improvement also correlated with improvements in quality of life for both groups, but more strongly for the Nasal Insert group. Thus, it is plausible that the use of nasal insert olfactory training leads to better subjective ability, which coincides with improved perceived quality of life.

Regarding more general physical and mental health related quality of life, only one subscale showed significant results of olfactory training. This is unsurprising. The SF-36 is a measure with an overall heavy focus on physical functioning, which arguably is less relevant for olfactory dysfunctions. Notwithstanding, the nasal insert group reported more improvement on the social functioning subscale compared to the standard care group at the post-training assessment. These results were somewhat surprising considering how we have previously concluded that one of the main drivers of lower quality of life amongst individuals with olfactory dysfunction is lack of enjoyment of food as well and worries related to coping with long-term dysfunctions, rather than problems related to interpersonal relationships (Anja L Winter et al., 2023). Interestingly, although not significant, the standard care group expressed nominally lower scores following the eight weeks of olfactory training than they did at baseline, contrary to the nasal insert group that expressed the opposite pattern. In addition, the difference between the groups was no longer evident after one year. One explanation for these findings could be the fact that standard care olfactory training is often viewed as tedious and time-consuming given that the hand-held delivery system used naturally comes with a greater restriction to the individuals’ mobility compared to the nasal inserts, which could affect participants’ view on how their problems interfere with social activities. In other words, these results may be attributable to diminished social functioning of the group training with the more tedious standard care protocol

One limitation of the present study is that when not using the LOFC method for the analyses, the group effects on subjective olfactory function (A L Winter et al., 2025) do not reach significance. However, considering the variation in sample sizes between the groups during the post training and 1-year follow-up measuring points (due to larger number of drop-outs in the standard care group) we argue that maintaining a consistent sample size allowed for a more complete comparison of effects between the groups while also reducing bias that could result from the exclusion of participants with incomplete data. If not, treatment conditions with lower drop-out rate are punished for their success. It is also reasonable to assume that using the last observed score for the individual provides a more conservative estimate, rather than exaggerating the effects of training, which replacement with the group mean (another common replacement method) might provide. For these reasons, we argue that the LOFC approach provides a more representative and informative depiction of the effects investigated.

Further, the study excluded many common olfactory dysfunction populations and used restricted inclusion criteria. The focus on the post-infectious sample may therefore have yielded results not directly generalizable to other etiologies of olfactory dysfunction. In addition, although olfactory dysfunction was assessed using standardized tests and common non-infectious causes were excluded, we cannot be certain that the dysfunction in all participants was truly post-infectious as this was not clinically verified.

Furthermore, due to feasibility reasons, the study protocol involved only eight weeks of olfactory training. This length of training is shorter than what is often described in the current worldwide literature on the topic, often stating a minimum of twelve weeks (Damm et al., 2014; Thomas Hummel et al., 2009). The optimal length of olfactory training is debated and not yet established; however, there are indications that longer treatment regimens produce better recovery (Altundag et al., 2015; Konstantinidis et al., 2016), likely due to the slow regeneration of olfactory receptor neurons. This fact could potentially conceal the potential effects of the olfactory training of this study had it been conducted using a more extensive regimen. In contrast, the required length of each training session in the present study was 20 minutes, which is longer compared to the training time often described in the current literature recommending smelling 4 odors for 10 seconds each, rendering a training session comprising around 40 seconds of effective training time repeated twice daily (Damm et al., 2014; Thomas Hummel et al., 2009). It could be argued that the longer odor exposure time during each session to some degree compensated for the shorter training time in weeks given that for all training, intensity and length of each session matters. The clinical effect of physical rehabilitation training is determined by multiple components of the exercise dose, including not only the overall duration of the regimen (weeks or months), but also the length and intensity of each training session. For example, randomized trials and meta-regression analyses in cardiac and stroke rehabilitation have shown that manipulating session time and exercise intensity at comparable regimen lengths alters clinical and physiological outcomes, which indicates that the relationship is not strictly linear (Abell et al., 2017; Sunderland et al., 1992). Future studies could therefore investigate the current topic of effects on quality of life and other subjective measures using variations in both session and regimen length.

In conclusion, this study demonstrates an overall long-lasting improvement of olfactory training on quality of life as well as subjective measures of qualitative and quantitative olfactory function. Conducting olfactory training using nasal inserts, rather than the standard regimen, also seems to yield a more robust improvement in subjective quantitative olfactory function as well as social functioning related quality of life.

## AUTHORSHIP CONTRIBUTION

JNL, ALW, and PSJ contributed to the conception and design of the study. ALW refined the study protocol, collected the data, performed the statistical analyses and wrote the first draft of the manuscript. JNL and PSJ wrote sections of the manuscript. JNL provided funding for the study. All authors edited versions, as well as read and approved the final version, of the manuscript.

## Data Availability

All raw data and analysis scripts are available at the Open Science Framework (OSF) https://osf.io/jtbf6/overview.

## REFERENCES

Abell, B., Glasziou, P., & Hoffmann, T. (2017). The Contribution of Individual Exercise Training Components to Clinical Outcomes in Randomised Controlled Trials of Cardiac Rehabilitation: A Systematic Review and Meta-regression. Sports Medicine - Open, 3(1), 19. 10.1186/s40798-017-0086-z

Altundag, A., Cayonu, M., Kayabasoglu, G., Salihoglu, M., Tekeli, H., Saglam, O., & Hummel, T. (2015). Modified olfactory training in patients with postinfectious olfactory loss. The Laryngoscope, 125(8), 1763–1766. 10.1002/lary.25245

Alwateer, M., Atlam, E.-S., El-Raouf, M. M. A., Ghoneim, O. A., & Gad, I. (2024). Missing data imputation: A comprehensive review. Journal of Computer and Communications, 12(11), 53–75. 10.4236/jcc.2024.1211004

Boesveldt, S., & Parma, V. (2021). The importance of the olfactory system in human well-being, through nutrition and social behavior. Cell and Tissue Research, 383(1), 559–567. 10.1007/s00441-020-03367-7

Brazier, J. E., Harper, R., Jones, N. M., O’Cathain, A., Thomas, K. J., Usherwood, T., & Westlake, L. (1992). Validating the SF-36 health survey questionnaire: new outcome measure for primary care. BMJ (Clinical Research Ed.), 305(6846), 160–164. 10.1136/bmj.305.6846.160

Croy, I., Nordin, S., & Hummel, T. (2014). Olfactory disorders and quality of life--an updated review. Chemical Senses, 39(3), 185–194. 10.1093/chemse/bjt072

Damm, M., Pikart, L. K., Reimann, H., Burkert, S., Göktas, Ö., Haxel, B., Frey, S., Charalampakis, I., Beule, A., Renner, B., Hummel, T., & Hüttenbrink, K.-B. (2014). Olfactory training is helpful in postinfectious olfactory loss: a randomized, controlled, multicenter study. The Laryngoscope, 124(4), 826–831. 10.1002/lary.24340

Digitale, J., Franzon, D., Pletcher, M. J., McCulloch, C. E., & Gennatas, E. D. (2025). Methods for addressing missingness in electronic health record data for clinical prediction models: comparative evaluation. JMIR Medical Informatics, 13, e79307. 10.2196/79307

DiMatteo, M. R., Giordani, P. J., Lepper, H. S., & Croghan, T. W. (2002). Patient adherence and medical treatment outcomes: a meta-analysis. Medical Care, 40(9), 794–811. 10.1097/01.MLR.0000024612.61915.2D

Hummel, T, Sekinger, B., Wolf, S. R., Pauli, E., & Kobal, G. (1997). “Sniffin” sticks’: olfactory performance assessed by the combined testing of odor identification, odor discrimination and olfactory threshold. Chemical Senses, 22(1), 39–52. 10.1093/chemse/22.1.39

Hummel, Thomas, Rissom, K., Reden, J., Hähner, A., Weidenbecher, M., & Hüttenbrink, K.-B. (2009). Effects of olfactory training in patients with olfactory loss. The Laryngoscope, 119(3), 496–499. 10.1002/lary.20101

Jaime-Lara, R. B., Parma, V., Yan, C. H., Faraji, F., & Joseph, P. V. (2020). Role of olfaction in human health: A focus on coronaviruses. Allergy & Rhinology (Providence, R.I.), 11, 2152656720928245. 10.1177/2152656720928245

Konstantinidis, I., Tsakiropoulou, E., & Constantinidis, J. (2016). Long term effects of olfactory training in patients with post-infectious olfactory loss. Rhinology, 54(2), 170–175. 10.4193/Rhino15.264

Lee, J. J., Mahadev, A., Kallogjeri, D., Peterson, A. M., Gupta, S., Khan, A. M., Jiramongkolchai, P., Schneider, J. S., & Piccirillo, J. F. (2022). Development and psychometric validation of the olfactory dysfunction outcomes rating. JAMA Otolaryngology--Head & Neck Surgery, 148(12), 1132–1139. 10.1001/jamaoto.2022.3299

R Core Team. (2024). *R: A language and environment for statistical computing. R Foundation for Statistical Computing* (4.3.3) [Computer software].

Seiden, A. M. (2004). Postviral olfactory loss. Otolaryngologic Clinics of North America, 37(6), 1159–1166. 10.1016/j.otc.2004.06.007

Senn, S. (2006). Change from baseline and analysis of covariance revisited. Statistics in Medicine, 25(24), 4334–4344. 10.1002/sim.2682

Sluijs, E. M., Kok, G. J., & van der Zee, J. (1993). Correlates of exercise compliance in physical therapy. Physical Therapy, 73(11), 771–782; discussion 783. 10.1093/ptj/73.11.771

Sunderland, A., Tinson, D. J., Bradley, E. L., Fletcher, D., Langton Hewer, R., & Wade, D. T. (1992). Enhanced physical therapy improves recovery of arm function after stroke. A randomised controlled trial. *Journal of Neurology*, Neurosurgery, and Psychiatry, 55(7), 530–535. 10.1136/jnnp.55.7.530

Ware, J. E., & Sherbourne, C. D. (1992). The MOS 36-item short-form health survey (SF-36). I. Conceptual framework and item selection. Medical Care, 30(6), 473–483.

Winter, A L, Henecke, S., Thunell, E., Swartz, M., Martinsen, J., Sahlstrand Johnson, P., & Lundstrom, J. N. (2025). Olfactory training using nasal inserts is more effective due to increased adherence. Rhinology, 63(4), 477–485. 10.4193/Rhin24.369

Winter, Anja L, Henecke, S., Lundström, J. N., & Thunell, E. (2023). Impairment of quality of life due to COVID-19-induced long-term olfactory dysfunction. Frontiers in Psychology, 14, 1165911. 10.3389/fpsyg.2023.1165911

Zimmermann, G., Pauly, M., & Bathke, A. C. (2019). Small-sample performance and underlying assumptions of a bootstrap-based inference method for a general analysis of covariance model with possibly heteroskedastic and nonnormal errors. Statistical Methods in Medical Research, 28(12), 3808–3821. 10.1177/0962280218817796

